# A prospective evaluation of inter-rater agreement of routine medical records audits at a large general hospital in São Paulo, Brazil

**DOI:** 10.1101/2020.01.21.20018325

**Authors:** Ana Carolina Cintra Nunes Mafra, João Luiz Miraglia, Fernando Antonio Basile Colugnati, Gilberto Soares Lourenço Padilha, Renata Rafaella Santos Tadeucci, Ederson Almeida, Mario Maia Bracco

**Affiliations:** Hospital Israelita Albert Einstein, São Paulo, Brasil; Hospital Municipal Dr. Moysés Deutsch – M’Boi Mirim, São Paulo, Brasil; Universidade Federal de Juiz de Fora, School of Medicine, Juiz de Fora, Brasil; Centro de Estudos e Pesquisas Dr. João Amorim – CEJAM, São Paulo, Brasil

**Author notes:** Corresponding author Avenida Brigadeiro Faria Lima, 1188, Jardim Paulistano, São Paulo – SP, Brasil CEP 01451-001.

**Keywords:** Inter-rater agreement, Longitudinal agreement, medical quality register, audit, Gwet’s AC1

## Abstract

**Background:** The quality of the patient’s medical records is strictly related to patient safety. Besides, its data are widely used in observational studies. However, the reliability of the information extracted from them is a matter of concern in audit processes to ensure inter-rater agreement (IRA). Thus, the objective of this study is to evaluate the IRA among members of the Patient’s Health Record Review Board (PHRRB), in routine auditing of medical records, and the impact of periodic discussions of results with raters.

**Methods:** Prospective longitudinal study conducted between July of 2015 and April of 2016 at Hospital Municipal Dr. Moysés Deutsch, a large public hospital in São Paulo. The PHRRB was composed of 12 physicians, 9 nurses and 3 physiotherapists, who audited medical records, monthly, with the number of raters changing throughout the study. It was carried out PHRRB meetings to reach a consensus on criteria that the members have to rate in the auditing process. It was created a review chart that raters should verify the registry of patient’s secondary diagnosis, chief complaint, history of presenting complaint, past medical history, medication history, physical exam and diagnostic testing. It was obtained the IRA every three months. The Gwet’s AC1 coefficient and Proportion of Agreement (PA) were calculated to evaluate the IRA for each item over time.

**Results:** The study included 1884 items from 239 records with an overall full agreement among raters of 71.2%. A significant IRA increase by 16.5% (OR=1.17; 95% CI=1.03—1.32; p=0.014) was found in the routine PHRRB auditing, with no significant differences between the PA and the Gwet’s AC1, that showed a similar evolution over time. The PA decreased by 27.1% when at least one of the raters was absent from the review meeting (OR=0.73; 95% CI=0.53—1.00; p=0.048).

**Conclusions:** Medical record quality has been associated with the quality of care and could be optimized and improved by targeted interventions. The PA and the Gwet’s AC1 are suitable agreement coefficients that are feasible to be incorporated in the routine of PHRRB evaluation process.

## Background

Adequate medical recordkeeping is an essential part of good health professional practice that makes it possible to evaluate and improve the quality of health care. Besides, medical records should also be used as a learning tool, far beyond the medical management of patients, but also improving the coordination and continuity of care and supporting decision making, avoiding adverse events that can compromise patient safety, mainly in hospitals.^1,2^

Auditing patient’s medical records is a practice that aims to ensure the quality of patient care throughout reliable information registered by health professionals during patient visits or admissions in healthcare units. The Brazilian Medical Council establishes that patients’ records review commissions is mandatory for health services, since 2002.^3^ However, the reliability of the auditing processes is a matter closely related to the inter-rater agreement (IRA) when different raters assign the same precise value for each item being observed^4,5^. Some review studies assessing adverse events have been shown to suffer from poor to moderate inter-rater reliability (IRR)^6,7^. Also, IRR is rarely described or discussed in research papers based on data abstracted from medical records and there are no standard methods for assessing IRR^8^. Moreover, time constraints and work overload are frequent situations faced by health staff to perform tasks involving data management resulting in low data quality that can affect managerial decision-making. Therefore, the evaluation of suitable methods for data abstraction from this source is essential.^9^

When such studies employ multiple raters it is important to have a strategy to document adequate levels of agreement between them, and the Cohen’s Kappa coefficient (κ) is a well-known measure.^10^ However, it is affected by the skewed distributions of categories (the prevalence paradox) and by the degree to which raters disagree (the bias problem).^11,12^

Kilem Li Gwet proposed a new agreement coefficient to fix those limitations and which can be used with any number of raters requiring a simple categorical rating system.^13,14^ The objective of this study was to evaluate the IRA of routine audits of medical records and the impact of periodic discussions among raters, to refine auditing criteria, in a large general hospital as part of an intervention to improve the quality of the medical records related to essential content. Besides, the study also aimed to compare the estimates of the percent agreement (PA) to the Gwet’s agreement coefficient (AC1) and to identify possible factors associated with the PA.

## Methods

### Population and setting

This was a prospective longitudinal study conducted between July of 2015 and April of 2016 at the Hospital Municipal Dr. Moysés Deutsch (HMMD), a large public general hospital (300 beds) located in the southern zone in the city of São Paulo, Brazil, an impoverished region encompassing approximately 600,000 inhabitants. The present study was part of a larger intervention aimed at improving the quality of patient care throughout a tailored integration strategies among health facilities in its Regional Health Care Network, that used a Lean Six Sigma methodology to get improvements of data quality registered in the medical records, with potential benefits to the patients, to decision-making actions and processes, and to obtain scientific quality over these data for research purposes, as published elsewhere.^15^

### Audit of medical records and review meetings

The HMMD maintains a routine auditing process that includes 13% of all medical records of patients discharged in the previous month, carried out by the Patient’s Medical Record Review Board (PMRRB), that was composed by 24 nominated health professionals from several HMMD staff, medical, nurses and multi-professional staff coordinators, or delegated by them, from all clinical departments, 12 physicians, 9 nurses and 3 physiotherapists. It is a time-costly procedure because competes with the patient-care tasks among these professionals. As a consequence, is common that the audits have happened isolated by each PMRRB member without any criteria alignment, to rate the items in the audit chart, which compromise the whole quality of the auditing process. However, the patient’s medical charts were selected in a non-aleatory way, lacking representativeness over the results achieved, compromising the accuracy and generalizability of these data.

The planned intervention has used the Lean-6-Sigma methodology, that is largely utilized to aggregate values in several HMMD quality improvement processes, that is part of the environment culture among the professionals.^15^

The proposed actions included at least one team-leader from each HMMD clinical department, preferably its coordinator, which increased the PMRRB components, reducing the total medical charts to be reviewed by each member. It was refined the audit chart by all members through discussions about the relevance of the information that should be registered by their health teams, answering the question: ‘Which information cannot be missed in the patient’s medical record?” The chosen items were then discussed, to define the criteria that should be rated as adequate, inadequate, incomplete, or not applicable (Table 1). Finally, the patient’s medical records have become selected randomly, weighed by the discharge proportion of each department.

**Table 1:** Audited items.

| **Audited items** | **Options to rate** |
| --- | --- |
| Secondary diagnosis | Adequate or inadequate |
| Chief complaint | Not applicable, adequate or inadequate |
| History of presenting complaint | Not applicable, adequate or inadequate |
| Past medical history | Not applicable, adequate or inadequate |
| Medication history | Not applicable, adequate or inadequate |
| Complete medical history | Not applicable, adequate, inadequate or incomplete |
| Physical exam | Adequate or inadequate |
| Diagnostic testing | Not applicable, adequate or inadequate |

The number of raters varying throughout the study as shown in Table 2.

**Table 2:** Number of audited medical records and raters over time.

| **Audit period** | **Number of medical**<br>**records for IRA** | **Number of raters** |
| --- | --- | --- |
| **1. 2015 July** | 54 | 18 |
| **2. 2015 October** | 45 | 19 |
| **3. 2016 January** | 84 | 21 |
| **4. 2016 April** | 56 | 16 |

Every three months during the study period, and in addition to the routine audits, five to six medical records were randomly allocated to the same two or three independent raters of the same professional category to evaluate the IRA. The study also included review meetings conducted every three months to align assessment criteria based on the results of the IRA evaluation and the auditing processes.

### Statistical analysis

The Gwet’s AC1 and PA were calculated to evaluate the IRA for each item over time and were compared through line graphs including 95% confidence intervals (CIs). The Gwet’s AC1 95% CIs^14^ were calculated, while the PA were modelled by generalized estimating equations (GEE), without an intercept^16,17^. The agreement measures were interpreted following the categories proposed by Altman.^18^

Logistic GEE was used to model the PA of all raters along the time, using the value of 1 to full agreement and of 0 to some disagreement, and combining all items for each item individually. The analyses employed an exchangeable working correlation matrix and items in a single medical record were considered to be correlated. The model included as independent variables: professional category, review meeting attendance, and time (audits 1 to 4). A forward stepwise approach was used to variable selection employing a p-value lesser than or equal to 0.200 in the unadjusted model, and lesser than or equal to 0.050 in the multiple-variable model.

The analyses were performed with the R software version 3.2.2^19^ with geepack^20^.

## Results

The study included 1884 items from 239 records with an overall full agreement among raters of 71.2%. The estimated mean PA was found to be larger than the Gwet’s AC1 for all audited items (Figure 1), however, these differences were not statistically significant and the evolution of the two agreement coefficients was similar throughout the study period. Although a positive trend was found in the agreement of almost all items, their CIs did not indicate any statistically significant changing over time. Also, the coefficients measurements got closer as the agreement got greater. During the study period, the greatest agreement was “chief complaint”, while the lowest one was “secondary diagnosis”.

**Figure 1:**
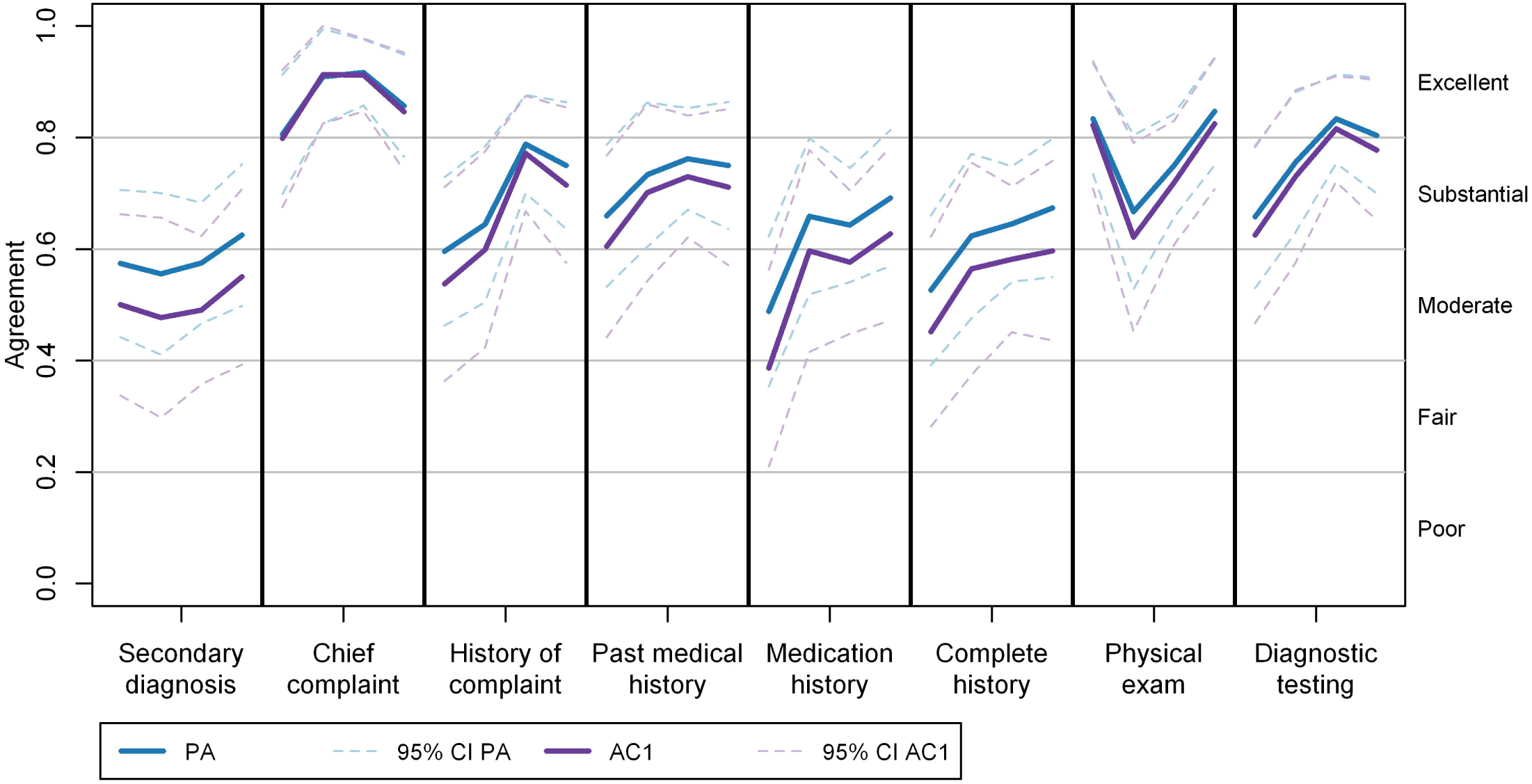
Estimated percent agreement (PA) and Gwet’s AC1 by audited item. CI: Confidence interval. AC: Agreement coefficient.

The logistic GEE model that included all items (Table 3) found a statistically significant increase of 17% over time for the PA, but when at least one of the raters was absent from the review meeting, the PA decreased by 27%. Physiotherapists and physicians showed higher PA when compared to nurses.

**Table 3:**
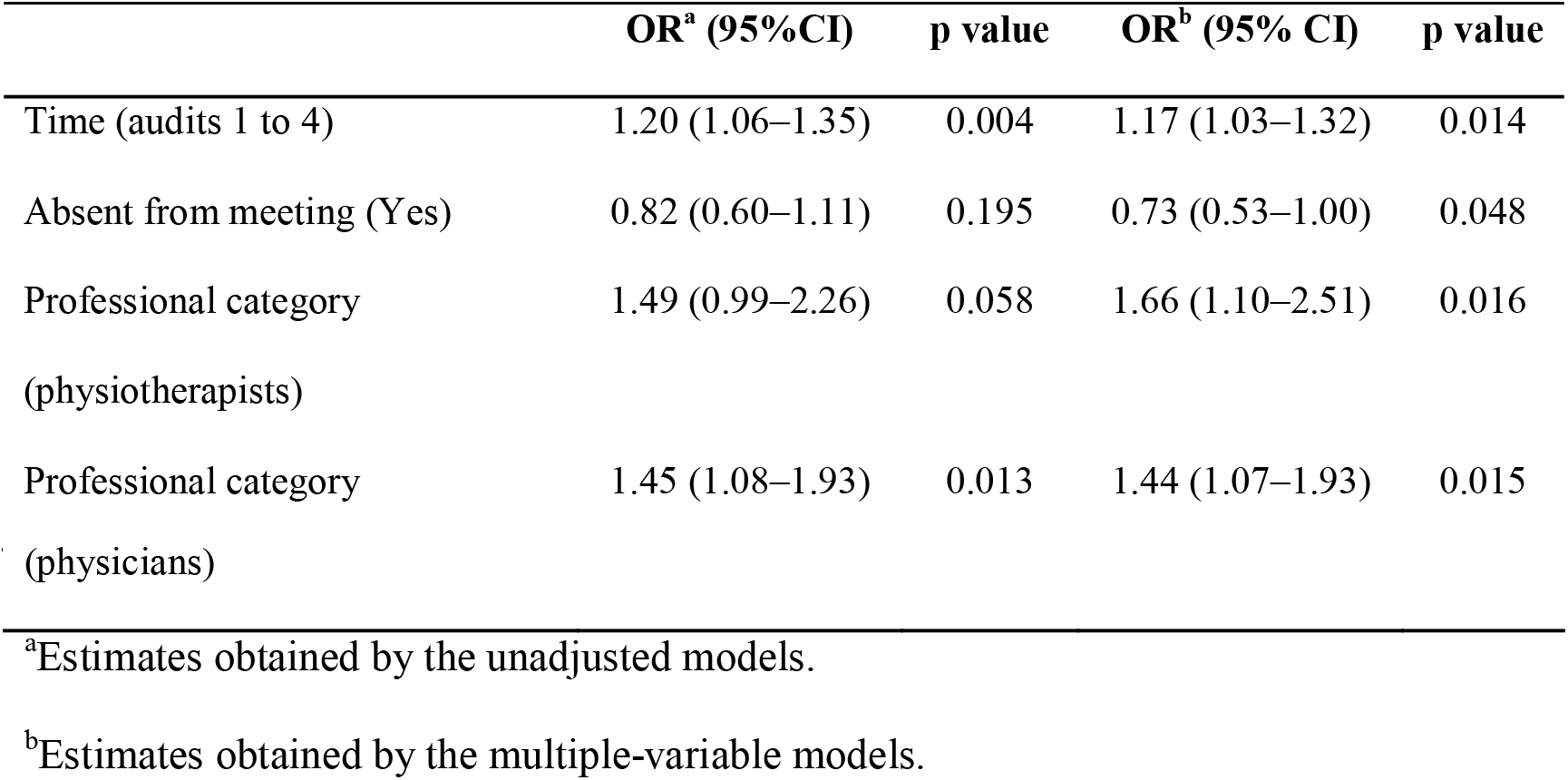
Estimated odds ratios (OR) for percent agreement. N=1884 items from 239

In the analysis by item, there was a non-significant positive trend to higher PA for History of presenting complaint while physicians presented a significantly higher PA over time when compared to nurses for “secondary diagnosis”, “medication history”, and “diagnostic testing”. Physiotherapists presented a significantly higher PA over time when compared to nurses for “medication history”. Finally, when at least one of the raters was absent from the review meeting the PA decreased by 60.5% for “diagnostic testing” (Table 4).

**Table 4:** Estimated odds ratios (OR) of percent agreement by item. N= 239 records.

|  | OR (95% CI) | p value |
| --- | --- | --- |
| Secondary diagnosis |  |  |
| Time (audits 1 to 4) | 1.02 (0.80–1.30) | 0.878 |
| Absent from meeting (Yes) | 0.65 (0.35–1.23) | 0.187 |
| Prof. category (physiotherapists) | 1.74 (0.72–4.22) | 0.221 |
| Prof. category (physicians) | 1.82 (1.00–3.29) | 0.048 |
| Chief complaint |  |  |
| Time (audits 1 to 4) | 1.20 (0.80–1.80) | 0.370 |

|  | **OR (95% CI)** | **p value** |
| --- | --- | --- |
| Absent from meeting (Yes) | 0.71 (0.26–1.95) | 0.507 |
| Prof. category (physiotherapists) | 1.22 (0.29–5.16) | 0.786 |
| Prof. category (physicians) | 0.74 (0.30–1.83) | 0.519 |
| History of presenting complaint |  |  |
| Time (audits 1 to 4) | 1.31 (0.99–1.74) | 0.056 |
| Absent from meeting (Yes) | 0.58 (0.28–1.19) | 0.138 |
| Prof. category (physiotherapists) | 1.78 (0.68–4.61) | 0.238 |
| Prof. category (physicians) | 1.56 (0.81–3.01) | 0.182 |
| Past medical history |  |  |
| Time (audits 1 to 4) | 1.14 (0.86–1.51) | 0.375 |
| Absent from meeting (Yes) | 0.70 (0.34–1.44) | 0.334 |
| Prof. category (physiotherapists) | 1.12 (0.43–2.91) | 0.817 |
| Prof. category (physicians) | 1.35 (0.69–2.63) | 0.378 |
| Medication history |  |  |
| Time (audits 1 to 4) | 1.25 (0.96–1.62) | 0.099 |
| Absent from meeting (Yes) | 0.83 (0.44–1.57) | 0.569 |
| Prof. category (physiotherapists) | 4.25 (1.53–11.77) | 0.005 |
| Prof. category (physicians) | 1.87 (1.02–3.41) | 0.041 |
| Complete medical history |  |  |
| Time (audits 1 to 4) | 1.22 (0.95–1.57) | 0.118 |
| Absent from meeting (Yes) | 0.74 (0.38–1.42) | 0.359 |
| Prof. category (physiotherapists) | 1.15 (0.47–2.82) | 0.753 |
| Prof. category (physicians) | 0.96 (0.52–1.76) | 0.893 |
| Physical exam |  |  |
| Time (audits 1 to 4) | 1.07 (0.82–1.39) | 0.616 |
| Absent from meeting (Yes) | 1.32 (0.68–2.58) | 0.410 |
| Prof. category (physiotherapists) | 2.59 (0.70–9.57) | 0.154 |
| Prof. category (physicians) | 1.02 (0.54–1.91) | 0.950 |
| Diagnostic testing |  |  |
| Time (audits 1 to 4) | 1.23 (0.91–1.66) | 0.175 |
| Absent from meeting (Yes) | 0.39 (0.18–0.89) | 0.024 |
| Prof. category (physiotherapists) | 1.52 (0.59–3.92) | 0.387 |
| Prof. category (physicians) | 3.11 (1.53–6.30) | 0.002 |

Prof.: professional.

## Discussion

A significant increase in the IRA among PHRRB members was found along the time in routine medical record auditing processes when periodic evaluations of the agreement were performed and discussed by them. On the other hand, but supporting this finding, the absence of a member in a review meeting had a negative impact in the PA. Besides, the PA and the Gwet’s AC1 were comparable and presented a similar evolution over time. Complete medical history was a composite of chief complaint, history of complaint, past medical history, and medication history. It was considered complete whether all of them were complete, too. Thus, it showed a positive evolution in both PA and Gwet AC1 over time from moderate to substantial according to Altman’s categories^18^. Only the IRA of secondary diagnosis has remained moderate. These findings can indicate raters learning curve regarding the positive evolution of some variables across agreement ranges. Nevertheless, the degree of agreement is arbitrary making it impossible to define an acceptable level^5^. Thus, the interpretation of these IRA values is following the main study objective, i.e., the rater’s concordance in a particular category.

The greater IRA among physicians and physiotherapists, when compared to nurses, can reflect some inconsistency over the evaluations that can be attributed by rater’s selection, training, and accountability^5^, that could be influenced by a misunderstanding about rating the Complete History item.

The strategy applied for the IRA was feasible to be carried out in this real-world scenario, aggregating value to the auditing process, providing more accurate information that can be used by health leadership. The use of PA and Gwet’s AC1 for that purpose was successful because they demand a relatively small sample of PMRs to be audited by each rater, and can provide two data consistency measures.^5,21^ Both of the used indices have reached acceptable levels of agreement^18,22^, according to study purposes.

Following and evaluating the progress of the agreement among raters of PMRs allows setting up goals and identifying associated factors to improve the audit processes, but previously proposed models worked with continuous variables^23^ or with the Kappa coefficient^24^, so the use of PA and Gwet AC1 made it possible to model the agreement of more than two raters over time.

The increased IRA highlights the need for more careful planning and evaluation of medical record audits since this activity is closely related to health care quality and patient safety improvements efforts.^8,9^

Since the present study was conducted under real-world conditions and included different health providers as raters, this intervention has the potential to be applicable in other similar settings, taking into consideration that it was carried out in only one hospital that has a culture of evidence-based improvement interventions, during a short-term follow-up. Furthermore, the literature on the quality of medical records keeping and IRA or IRR is scarce what is reflected by the fact that no reviews on the subject could be identified making the results of this study relevant to improve the body of knowledge, in the era of data-driven institutions and big data from patient’s health records.

Finally, this study did not include an evaluation of the impact in the quality of medical records what should be the final goal of any routine audit, and therefore should be evaluated in future studies.

## Data Availability

The dataset supporting the conclusions of this article is available to researchers who want to explore the data. To request, please send an email to.

## Abbreviations

AC1: agreement coefficient
CI: confidence interval
GEE: generalized estimating equations
HMMD: Hospital Municipal Dr. Moysés Deutsch
IRA: inter-rater agreement
IRR: inter-rater reliability
OR: odds ratio
PA: proportion of agreement
PHRRB: patient’s health record review board

## Declarations

### Ethics approval

The study was approved by the research ethics committee of the São Paulo Municipal Health Department, and the partners’ institutions (26981514.3.0000.0086).^25^

### Consent for publication

Not applicable.

### Competing interests

The authors declare that they have no competing interests.

### Funding

This work was funded by the Brazilian Ministry of Health and São Paulo State Research Foundation through Research Program for the Unified Health System-PPSUS grant 2012/51228-9.

### Author’s contribution

ACCNM conceptualized the study design, drafted the initial manuscript, carried out the random sampling, the statistical analysis, and revised the manuscript. RRST and EA elaborated and operationalized the intervention, contributed to the study design and reviewed the manuscript. FABC contributed to the study design and reviewed the manuscript. JLM and GP contributed to the interpretation of data for the work and revised the manuscript critically for important intellectual content. MMB elaborated the study design, operationalized the intervention, drafted and revised the manuscript. All authors approved the final manuscript as submitted.

## Acknowledgements

We thank all the MRRC members who conducted the audit records and support the process, the members of the archiving sector as well as the hospital leadership.

## Notes

### Competing Interest Statement

The authors have declared no competing interest.

### Funding Statement

This work was funded by the Brazilian Ministry of Health and Sao Paulo State Research Foundation (FAPESP) through Research Program for the Unified Health System-PPSUS grant 2012/51228-9.

